# Associations between biomarkers and p-wave indices in relation to atrial fibrillation development in heart failure patients

**DOI:** 10.1101/2025.06.24.25330237

**Authors:** Z. Nezami, A. Jujic, Marcus Ohlsson, M. Magnusson, H. Holm Isholth, P. G. Platonov

## Abstract

**Background:** The predictive value of atrial conduction abnormalities, reflected by P-wave indices (PWI), and their association with biomarkers signaling fibrosis for the development of atrial fibrillation (AF) in patients with heart failure (HF) remains underexplored. To address this gap, we investigated the associations between PWI, fibrotic biomarkers, and the risk of incident AF in a cohort of HF patients.

**Methods:** A total of 475 patients with new-onset or worsening HF, were followed for 5 years. Fibrosis-associated biomarkers (TIMP-2, MMP-2, MMP-3, MMP-9, ST-2, GDF-15, Gal-3) were analyzed using proximity extension assay. PWI (P-wave duration, P-wave amplitude in lead I, P-wave terminal force in V1, P-wave axis and P-wave morphology in inferior leads required for definition of interatrial block (IAB)) were derived from ECGs processed with the Glasgow algorithm. Cox regression assessed associations between the biomarkers, PWI, and incident AF.

**Results:** Among 475 individuals (mean age 74.6 years; 68% male), 41 developed incident AF over 5 years. Low P-wave amplitude at inclusion correlated negatively with GDF-15 (p < 0.001) and MMP-2 (p = 0.037) in both leads I and II. Six biomarkers were significantly associated with incident AF in adjusted analysis: TIMP-4 (HR 2.06, p=0.007), MMP-2 (HR 2.09, p=0.046), MMP-3 (HR 1.48, p=0.007), ST-2 (HR 1.66, p=0.003), GDF-15 (HR 2.26, p=0.001), and Gal-3 (HR 2.14, p=0.048). Among the PWI, P-wave axis <0° (HR: 4.74, p = 0.021) and low P-wave amplitude in lead I (HR: 2.09, p=0.036) were significantly associated with incident AF.

**Conclusions:** In this long-term prospective follow-up study of patients admitted for HF, biomarkers with proven associations to fibrosis were associated with incident AF. This study also showed that low P-wave amplitude may reflect abnormal LA-conduction pathway and displaced intra-atrial conduction pattern in advanced HF, as low P-wave in lead I and abnormal P-wave axis (<0 degree) were associated with incident AF.

## Introduction

Structural and electrophysiological atrial abnormalities are closely associated with an increased risk of atrial fibrillation (AF), often serving as the earliest clinical indication of atrial cardiomyopathy[1]. These abnormalities reflect underlying processes such as atrial remodeling, fibrosis, and conduction disturbances, which can be effectively captured through non-invasive measures. Among these, P-wave indices (PWI) on surface electrocardiography have gained prominence as reliable markers of atrial cardiomyopathy[2, 3]. These indices include P-wave duration (PWD), P-wave terminal force in lead V1 (PTFV1), abnormal P-wave axis (P-axis), and interatrial block (IAB) [2, 3], each providing unique insights into atrial electrical and structural integrity. Epidemiological studies have consistently demonstrated that prolonged PWD and increased PTFV1 are strongly associated with the onset of AF [4, 5]. Further, advanced IAB contributes to interatrial desynchrony and electromechanical dysfunction, and is often observed in the presence of atrial dilation, elevated atrial pressure, and fibrosis - key substrates for AF development [6]. As AF frequently coexists with heart failure (HF), it presents considerable clinical challenges, with each condition facilitating the occurrence and progression of the other [7]. The prevalence of AF among patients with HF varies widely, ranging from 23% to 65% depending on HF severity and etiology, and has repeatedly been shown to worsen prognosis by increasing the risk of hospitalizations, morbidity, and mortality [7, 8]. Therefore, preventing its onset in patients with HF has emerged as a critical therapeutic goal. Despite this, research on the prognostic utility of PWI for predicting incident AF in the acute setting of HF, where advanced atrial remodeling is common, remains limited. Both AF and HF share overlapping pathophysiological mechanisms, including structural remodeling, myocardial fibrosis, and neurohormonal activation. These interconnected processes not only drive disease progression but also underscore the bidirectional relationship between these conditions. In previous work, we identified myocardial fibrosis as a potential common denominator linking HF and AF [9], demonstrating a strong association between circulating biomarkers of myocardial fibrosis and prevalent AF [10]. Building on these findings, the present study aims to investigate the association between PWI and biomarkers of myocardial fibrosis in relation to incident AF among patients with HF.

## Methods

### Study population

The HeARt and Brain Falure inVESTigation study (HARVEST) is a prospective study conducted at Skane University Hospital, Sweden, focusing on patients hospitalized for the diagnosis of HF (ICD-10: I50-) [10]. Patients admitted to internal medicine or cardiology department for the treatment of newly diagnosed or exacerbated chronic HF are included, excluding those unable to provide informed consent, with no additional exclusion criteria applied. Between 20th of March 2014 and 1st of November 2021, a total of 475 with new-onset or worsening of HF were prospectively recruited in the observational study and followed-up for a maximum of 5 years.

The date of AF debut was ascertained by either the earliest documentation of the ICD-10 code I48 in the medical records or the first ECG demonstrating AF in the regional digital ECG database (MUSE, GE Healthcare, Milwaukee, WI, USA). Individuals diagnosed with AF prior to inclusion (either permanent or paroxysmal/persistent) were classified as having prevalent AF. Those who were in sinus rhythm at baseline but exhibited AF on ECG prior to inclusion were classified as having paroxysmal AF. Individuals who consistently showed no ECG evidence of AF prior to enrollment or at enrolment were categorized as having no history of AF and were included in analyses of incident AF. Patient group allocation and data availability are illustrated in **Figure 1**.

**Figure 1.**
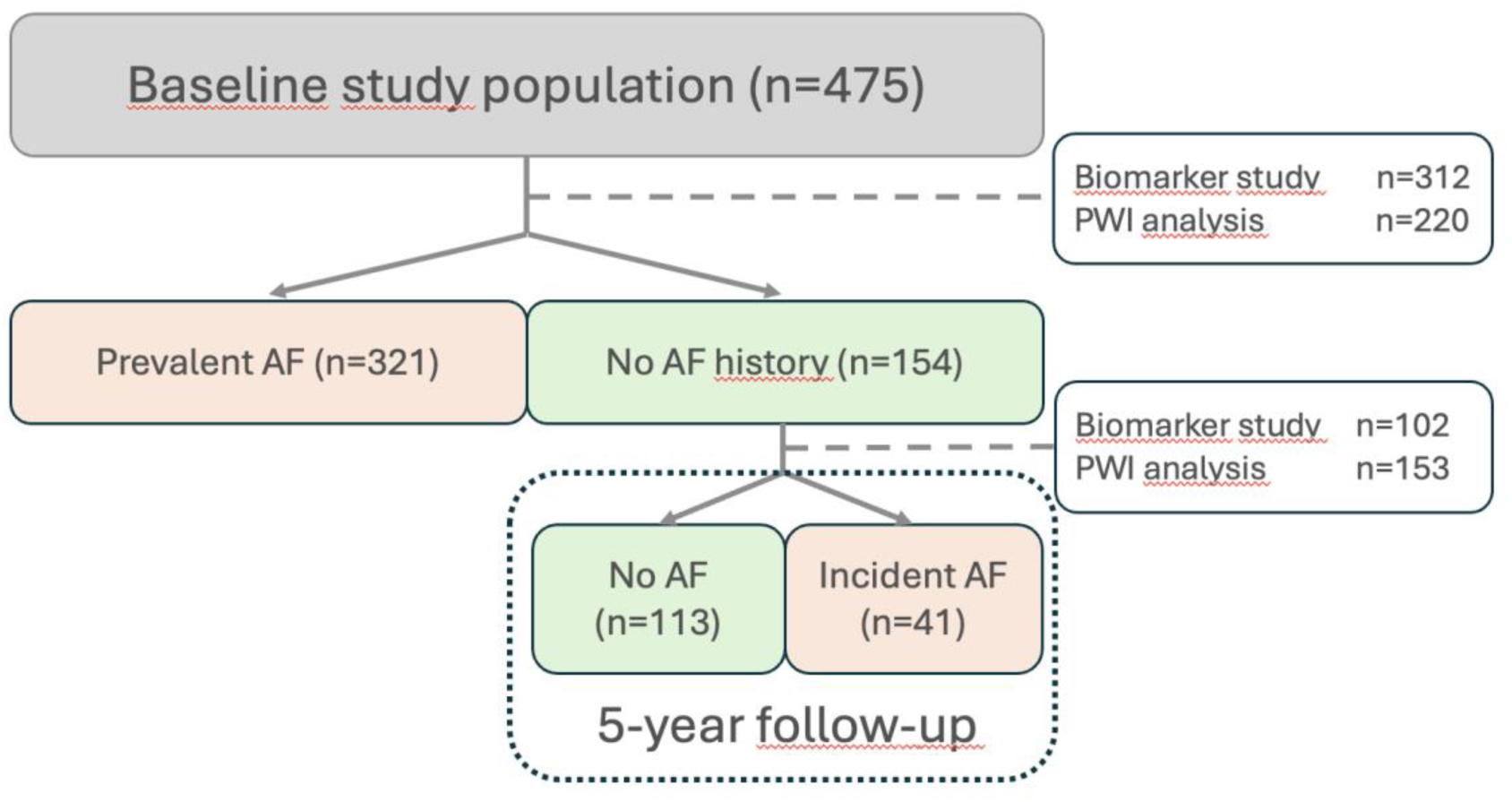
Flowchart of study population and data availability. Follow up time = 5 years.

The Ethical Review Board at Lund University, Sweden has approved the study, which complied with the Declaration of Helsinki. Written informed consent was obtained from all participants or relatives as described above.

### P-wave indices

All ECGs ever recorded in the hospital catchment area were exported in digital format retrieved from an electronic database (MUSE, GE Healthcare, Milwaukee, WI, USA) and processed offline using the Glasgow algorithm for calculation of the PWI (P-wave duration [PWD], P-wave amplitude in lead I, P-wave terminal force in V1 [PTFV1] and P-wave axis [P-axis]) and rhythm identification. PTFV1 of >0.04 mm×ms as being abnormal[11]. A P-wave axis <0◦ and >75◦ was considered abnormal [2]. A P wave duration (PWD) ≥120 milliseconds (ms) is defined as abnormal [12]. Interatrial block (IAB) is defined as partial if PWD > 120 ms and positive P-wave morphology in leads aVF and II or advanced if PWD exceeds 120 ms and P-wave is biphasic lead II and/or aVF [13]. ECGs with non-sinus rhythm were excluded from the analysis after manual ECG review (ZN).

### Proteomic profiling

Plasma levels of 92 cardiovascular related proteins (CVD) were measured using the Proseek Multiplex CVD III 96 x 96 kit (Olink Bioscience, Sweden) and the proximity extension assay (PEA) technique in 324 consecutive patients between March 2013 and January 2018. This method uses pairs antibodies labeled with DNA strands that binds to each target protein, allowing a unique polymerase chain reaction sequence to form and be quantified. All data are presented as arbitrary units. The CVD III panel includes 92 proteins, linked to metabolism, inflammation, and cardiovascular disease (CVD). The average variability was within assays 8.1% and 11.4%, respectively. Additional information regarding the assays is available on the Olink homepage. Seven proteins included in the CVD III panel were selected to be part of further analyses. The selected proteins have been previously associated with myocardial fibrosis including, metalloproteinase inhibitor 4 (TIMP-4) [14], suppression of tumorigenicity 2 (ST-2) [15], galectin-3 (Gal-3) [16], growth/differentiation factor-15 (GDF-15) [17], and matrix metalloproteinase 2, 3, and 9 (MMP-2, MMP-3, and MMP-9, respectively) [18, 19].

### Co-variates

Anthropometric measurements and blood samples were collected from patients after an overnight fast. Blood pressures were measured by trained nurses using a validated automated BP monitor Boso Medicus (Bosch + Sohn GmbH u. Co. KG, Jungingen, Germany). Hypertension was defined as either systolic blood pressure (SBP) ≥140 mmHg and/or diastolic blood pressure (DBP) ≥90 mmHg. Body mass index (BMI) was determined as weight in kilograms divided by height in meters squared, and information on participants’ medication use was recorded. Diabetes was classified as either a self-reported diagnosis of type 2 diabetes, current use of antidiabetic medications, or fasting plasma glucose levels exceeding 7 mmol/L. Smoking status was categorized based on self-report as either smoker or non-smoker; individuals who had never smoked were classified as non-smokers, while those who currently smoked were categorized as smokers. Analyses of total N-terminal prohormone BNP (NT-proBNP), creatinine, and additional biomarkers were conducted at the Department of Clinical Chemistry, Skåne University Hospital in Malmö, which adheres to the national standardization and quality assurance program (EQUALIS).

### Statistical analyses

Continuous variables are presented as means with standard deviations (SD) or as medians with interquartile ranges (25th–75th percentile), depending on their distribution. Categorical variables are summarized as counts and percentages. Group comparisons for continuous variables were performed using one-way analysis of variance (ANOVA) for normally distributed data and the Kruskal–Walli’s test for non-normally distributed data. For categorical variables, Pearson’s chi-square test was used to assess differences between groups. Logistic regression analyses were conducted to assess the cross-sectional associations between biomarkers, PWI, and paroxysmal AF. For the biomarker association with paroxysmal AF, adjusted p-values were calculated using the Benjamini–Hochberg method to control for multiple testing. A 5% false discovery rate (FDR) identified proteins for further analysis in Model 1 (adjusted for age and sex). Significant associations were analysed in Model 2 (adjusted for age, sex, systolic BP and NT-proBNP).

Follow-up time was defined as the interval from study enrollment to the earliest of (1) first AF diagnosis, (2) death, (3) censoring (completion of the five-year observation period), treating death as non-informative censoring event.

To evaluate predictors of incident AF, hazard ratios (HR) with 95% confidence intervals (CI) were calculated using Cox proportional hazards regression models. Adjustments were made in two models: Model 1 included age and sex, while Model 2 further adjusted for age, sex, NT-proBNP, and systolic blood pressure. The proportional hazards assumption was checked using Schoenfeld residuals. Kaplan-Meier curves were used to illustrate the associations.

A nominal *P*-value of < 0.05 was considered statistically significant. Unadjusted and adjusted logistic regressions for associations of fibrotic proteins with prevalent AF were performed.

Spearman’s correlation analysis was used to assess the correlation between PWI and biomarkers. It calculates a correlation coefficient (ρ or *rₛ*) ranging from-1 to +1, where: +1 indicates a perfect positive correlation and,-1 indicates a perfect negative monotonic relationship. 0, indicates no monotonic relationship. All statistical analyses were performed using the SPSS statistics software, version 27 for Mac, and R version 4.4.3, using the ggplot2-package.

## Results

### Baseline patient characteristics

Clinical and demographical characteristics of the study cohort are presented in **Table 1**. Median follow-up time was 3.3 (1.2-5.0) years. Compared to patients with prevalent AF at baseline (n=321), individuals without a history of AF (n=154) were younger and exhibited higher systolic-and diastolic blood pressure. This group were more likely to be current smokers and demonstrated better renal function, with higher eGFR values. Patients without AF history more frequently received ACE inhibitors. On ECG, participants without history of AF showed a lower prevalence of abnormal P-axis orientation (<0°: 4% vs 13%, >75°: 14% vs 27%), higher P-wave amplitude in lead I and a lower occurrence of advance IAB (3% vs 15%), as shown in **Table 1**. Based on these findings, we focused our analysis on P-wave amplitude in lead 1 to evaluate its prognostic significance in patients without prior history of AF, using tertile-based stratification.

**Table 1.**
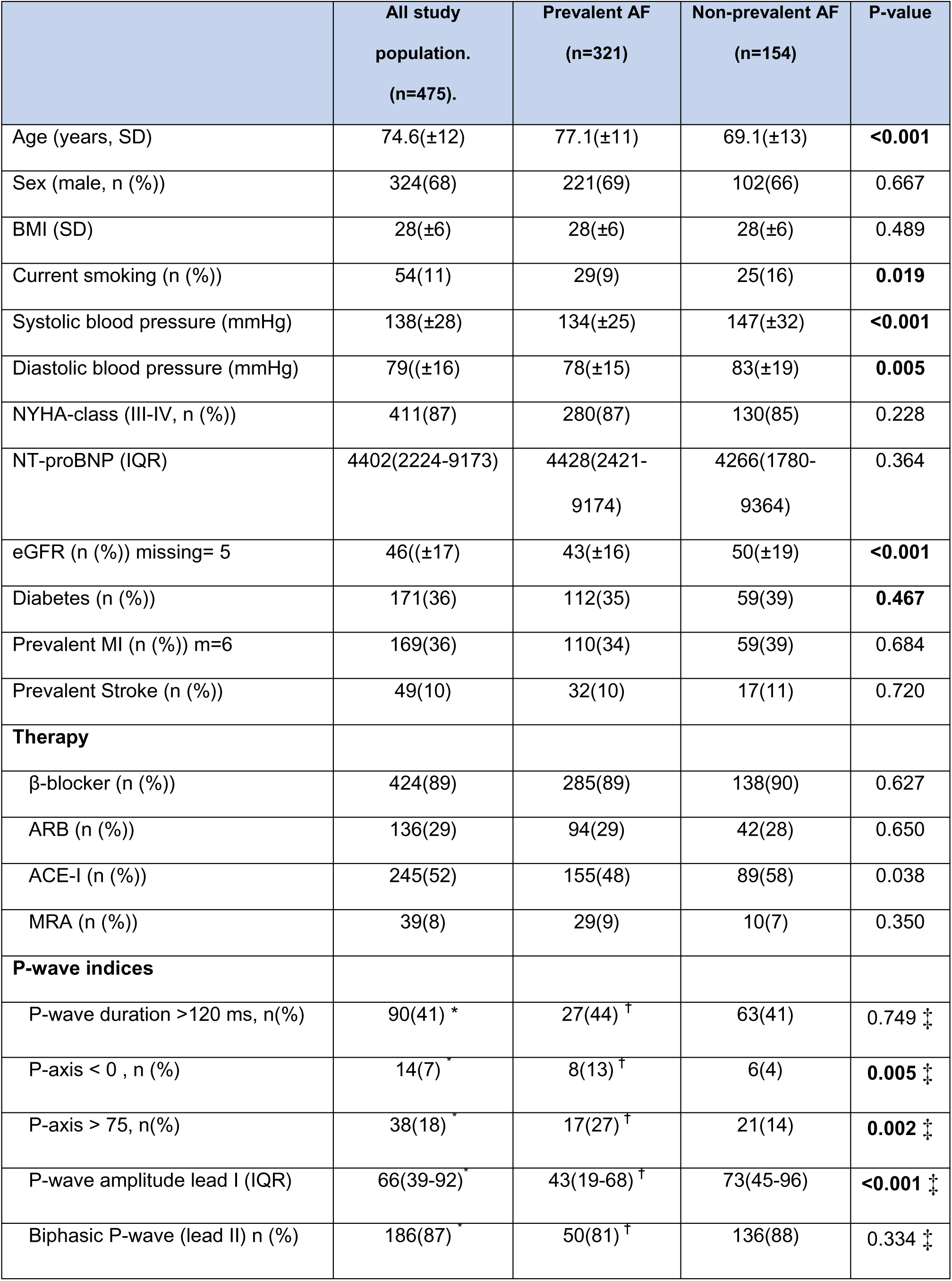
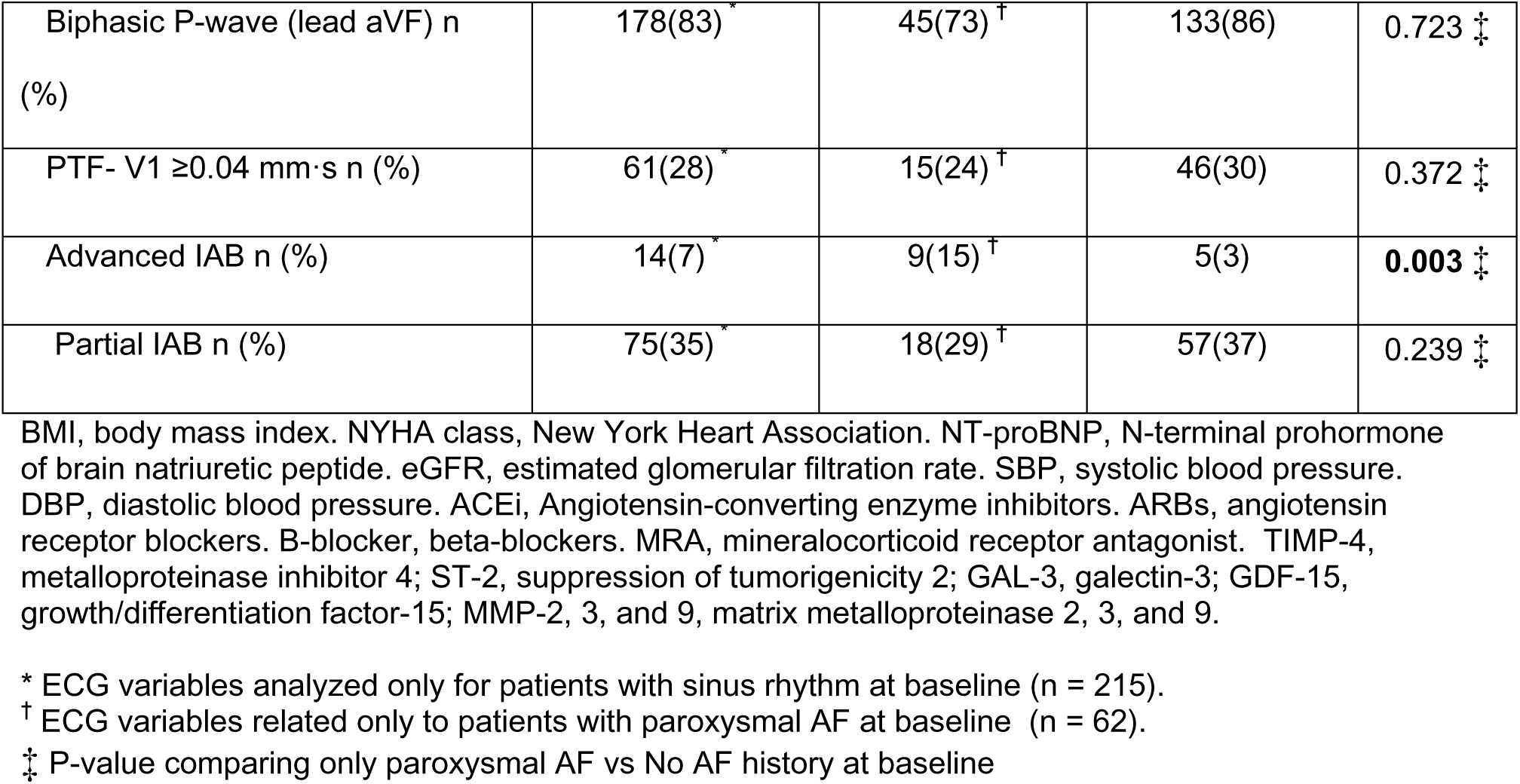
Baseline characteristics and PWI across AF groups.

|  | All study<br>population.<br>(n=475). | Prevalent AF<br>(n=321) | Non-prevalent AF<br>(n=154) | P-value |
| --- | --- | --- | --- | --- |
| Age (years, SD) | 74.6(±12) | 77.1(±11) | 69.1(±13) | **<0.001** |
| Sex (male, n (%)) | 324(68) | 221(69) | 102(66) | 0.667 |
| BMI (SD) | 28(±6) | 28(±6) | 28(±6) | 0.489 |
| Current smoking (n (%)) | 54(11) | 29(9) | 25(16) | **0.019** |
| Systolic blood pressure (mmHg) | 138(±28) | 134(±25) | 147(±32) | **<0.001** |
| Diastolic blood pressure (mmHg) | 79(±16) | 78(±15) | 83(±19) | **0.005** |
| NYHA-class (III-IV, n (%)) | 411(87) | 280(87) | 130(85) | 0.228 |
| NT-proBNP (IQR) | 4402(2224-9173) | 4428(2421-9174) | 4266(1780-9364) | 0.364 |
| eGFR (n (%)) missing= 5 | 46(±17) | 43(±16) | 50(±19) | **<0.001** |
| Diabetes (n (%)) | 171(36) | 112(35) | 59(39) | **0.467** |
| Prevalent MI (n (%)) m=6 | 169(36) | 110(34) | 59(39) | 0.684 |
| Prevalent Stroke (n (%)) | 49(10) | 32(10) | 17(11) | 0.720 |
| **Therapy** |  |  |  |  |
| β-blocker (n (%)) | 424(89) | 285(89) | 138(90) | 0.627 |
| ARB (n (%)) | 136(29) | 94(29) | 42(28) | 0.650 |
| ACE-I (n (%)) | 245(52) | 155(48) | 89(58) | 0.038 |
| MRA (n (%)) | 39(8) | 29(9) | 10(7) | 0.350 |
| **P-wave indices** |  |  |  |  |
| P-wave duration >120 ms, n(%) | 90(41) * | 27(44) † | 63(41) | 0.749 ‡ |
| P-axis < 0°, n (%) | 14(7) * | 8(13) † | 6(4) | **0.005** ‡ |
| P-axis > 75°, n(%) | 38(18) * | 17(27) † | 21(14) | **0.002** ‡ |
| P-wave amplitude lead I (IQR) | 66(39-92) * | 43(19-68) † | 73(45-96) | **<0.001** ‡ |
| Biphasic P-wave (lead II) n (%) | 186(87) * | 50(81) † | 136(88) | 0.334 ‡ |
| Biphasic P-wave (lead aVF) n (%) | 178(83)<sup>*</sup> | 45(73)<sup>†</sup> | 133(86) | 0.723<sup>‡</sup> |
| PTF- V1 ≥0.04 mm·s n (%) | 61(28)<sup>*</sup> | 15(24)<sup>†</sup> | 46(30) | 0.372<sup>‡</sup> |
| Advanced IAB n (%) | 14(7)<sup>*</sup> | 9(15)<sup>†</sup> | 5(3) | **0.003<sup>‡</sup>** |
| Partial IAB n (%) | 75(35)<sup>*</sup> | 18(29)<sup>†</sup> | 57(37) | 0.239<sup>‡</sup> |

BMI, body mass index. NYHA class, New York Heart Association. NT-proBNP, N-terminal prohormone of brain natriuretic peptide. eGFR, estimated glomerular filtration rate. SBP, systolic blood pressure. DBP, diastolic blood pressure. ACEi, Angiotensin-converting enzyme inhibitors. ARBs, angiotensin receptor blockers. B-blocker, beta-blockers. MRA, mineralocorticoid receptor antagonist. TIMP-4, metalloproteinase inhibitor 4; ST-2, suppression of tumorigenicity 2; GAL-3, galectin-3; GDF-15, growth/differentiation factor-15; MMP-2, 3, and 9, matrix metalloproteinase 2, 3, and 9.
\* ECG variables analyzed only for patients with sinus rhythm at baseline (n = 215).
† ECG variables related only to patients with paroxysmal AF at baseline (n = 62).
‡ P-value comparing only paroxysmal AF vs No AF history at baseline

### Association between biomarkers, P-wave indices, and prevalent atrial fibrillation

In the multivariable logistic regression analysis, elevated plasma levels of TIMP-4, MPP-2, and GDF-15 were significantly associated with higher likelihood of prevalent AF at baseline. In contrast, higher plasma levels of MPP-9 were less likely to have documented AF by baseline in the unadjusted model, **Table 2**. Analysis of correlation of PWI with fibrosis biomarkers at inclusion in the study demonstrated that low P-wave amplitude significantly correlated with GDF-15 (Spearmans’rho-0.335, p<0.001 for lead I), and MMP-2 (Spearman’s rho-0.178, p=0.037 for lead I). MMP-9 on the other hand had a positive correlation (Spearman’s rho 0.178, p=0.037 for lead I). The rest of the biomarkers did not show significant correlations with PWI, **Figure 3**.

**Table 2.** Logistic regression analyses for associations between fibrotic biomarkers and prevalent AF at baseline (n=312, including 210 patients with prevalent AF at baseline).

| Biomarker | Model 1 |  |  | Model 2 |  |  | Model 3 |  |  |
| --- | --- | --- | --- | --- | --- | --- | --- | --- | --- |
|  | OR | 95%CI | P-value | OR | 95%CI | P-value | OR | 95%CI | P-value |
| TIMP-4 | **2.30** | **1.55-3.40** | **<0.001** | **1.90** | **1.24-2.89** | **0.003** | **1.85** | **1.12-2.86** | **0.006** |
| MMP-2 | **2.05** | **1.35-3.11** | **<0.001** | **1.98** | **1.27-3.07** | **0.003** | **1.77** | **1.11-2.82** | **0.017** |
| MMP-3 | 1.25 | 0.98-1.59 | 0.080 | 1.05 | 0.81-1.37 | 0.724 | 1.01 | 0.76-1.33 | 0.970 |
| MMP-9 | **0.74** | **0.58-0.94** | **0.013** | 0.79 | 0.61-1.01 | 0.063 | 0.78 | 0.60-1.01 | 0.062 |
| ST-2 | **1.38** | **1.02-1.80** | **0.019** | **1.32** | **1.00-1.75** | **0.048** | 1.26 | 0.93-1.72 | 0.139 |
| GDF-15 | **1.84** | **1.38-2.46** | **<0.001** | **1.51** | **1.13-2.03** | **0.006** | **1.48** | **1.07-2.05** | **0.019** |
| Gal-3 | 1.31 | 0.83-2.07 | 0.250 | 1.03 | 0.64-1.67 | 0.902 | 0.92 | 0.56-1.49 | 0.729 |

TIMP-4, Metalloproteinase inhibitor 4. Matrix metalloproteinase 2, 3, and 9 (MMP-2, MMP-3, and MMP-9, respectively). ST-2, suppression of tumorigenicity 2. GDF-15. Growth/differentiation factor 15. GAL-3, galectin-3.
OR: Odds ratio
CI: Confidence interval
Model 1: Unadjusted
Model 1: Adjusted for Age, Sex
Model 2: Adjusted for Age, Sex, systolic blood pressure, and NT-proBNP

### Association between biomarkers, P-wave indices and incident AF

Of the 154 with patients who did not have AF history by enrollment in the study, 41 (8%) had AF newly diagnosed during 5-year follow-up.

Six biomarkers showed significant associations with incident AF in adjusted Cox-regression analyses: TIMP-4, MMP-2, MMP-3, ST-2, GDF-15 and Gal-3, **Table 3 and Figure 2**. Similarly to the cross-sectional logistic regression analysis performed at baseline, PWI, particularly P-wave axis <0° and low P-wave amplitude in lead I demonstrated strong associations with incident AF in the adjusted Cox regression analysis, **Table 4 and Figure 4**.

**Figure 2.**
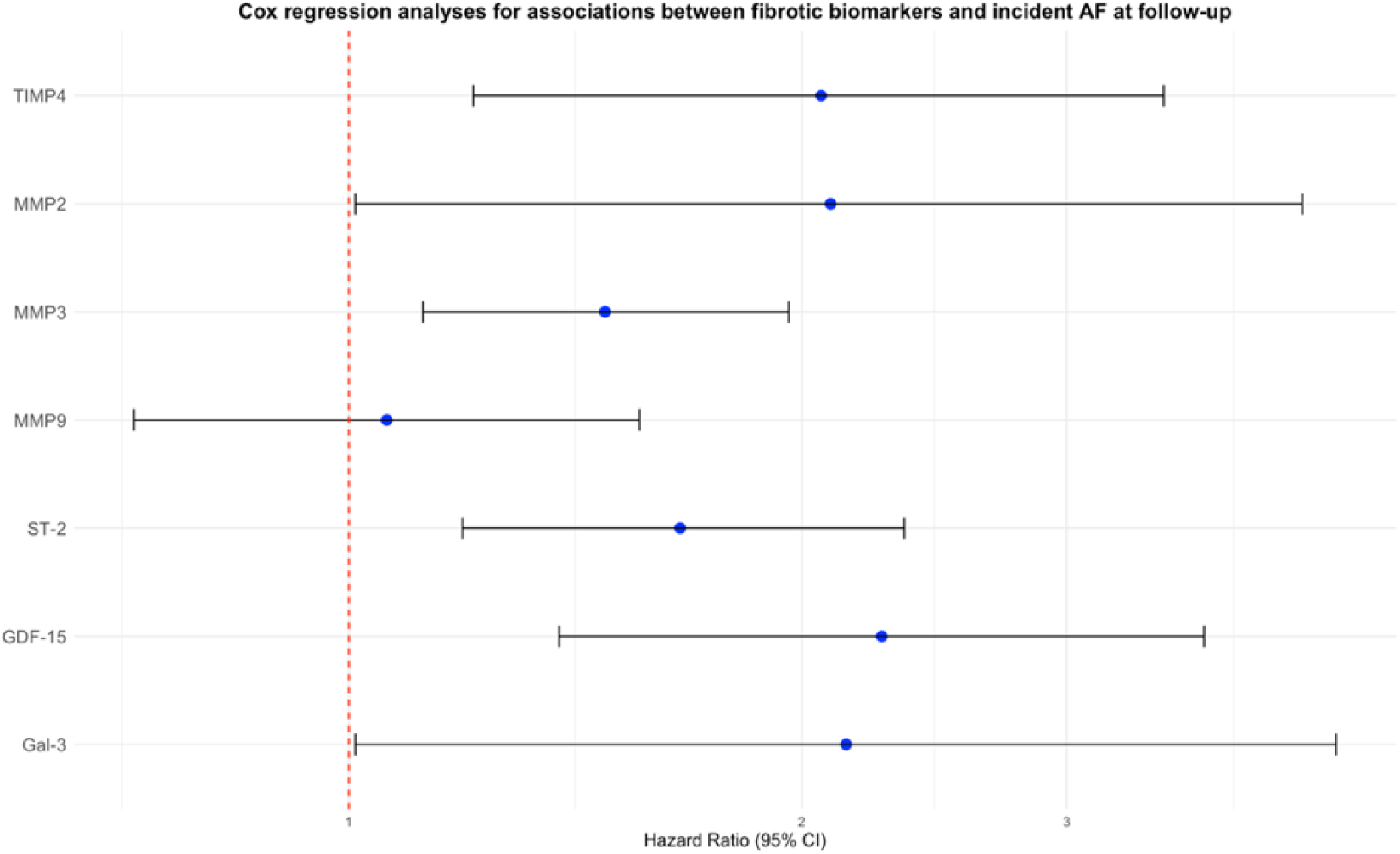
Forest plot illustrating association between the selected fibrosis-associated biomarkers and incident AF in patients with heart failure enrolled in the HARVEST study. Cox regression analysis was performed with adjustment for age, sex, systolic blood pressure and NT-proBNP.

**Figure 3.**
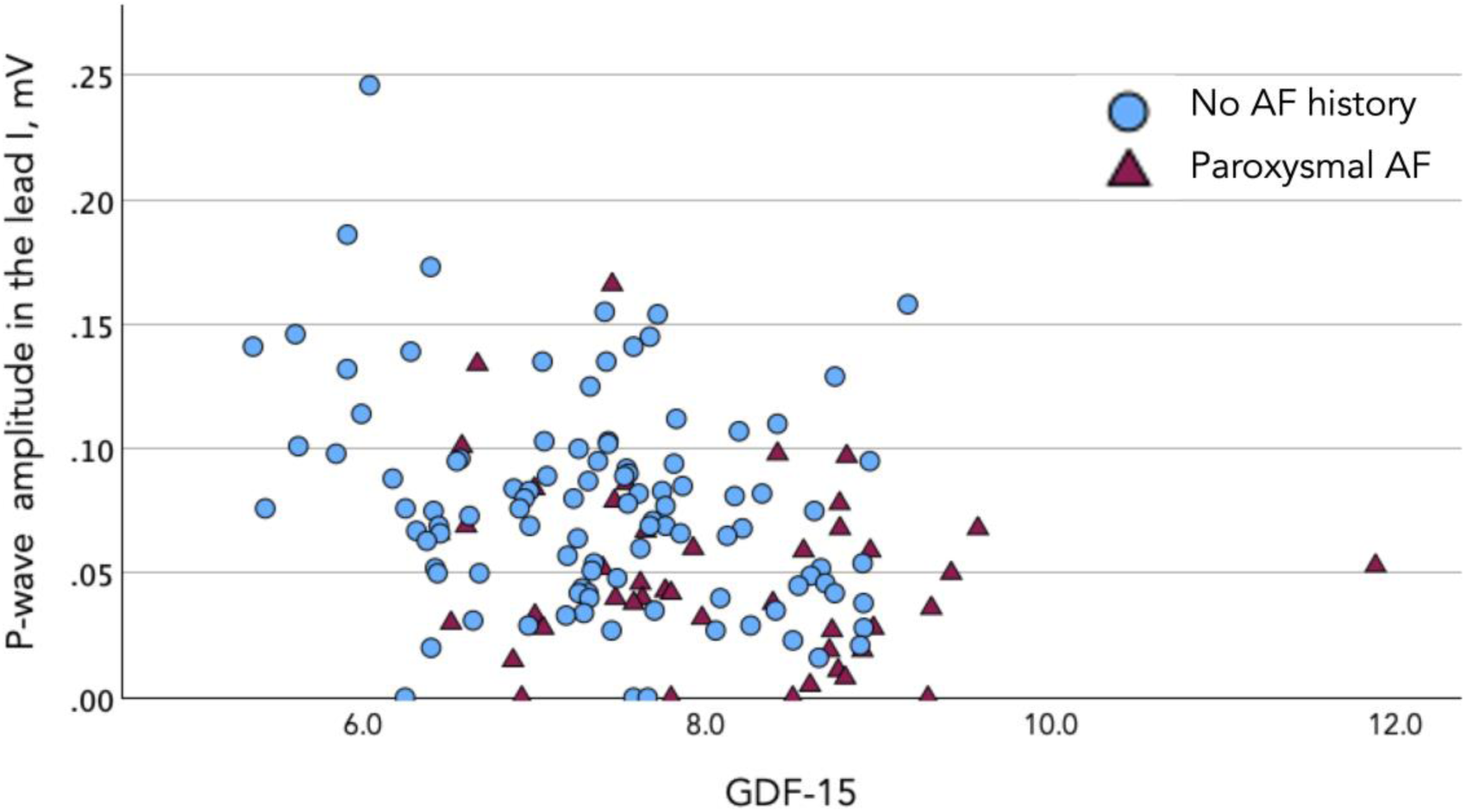
Scatter plot illustrating the correlation between P-wave amplitude in lead I at baseline ECG and GDF-15. GDF-15, which has been associated with fibrosis is inversely related to the P-wave amplitude ( Spearman’s *r=*-0.335, p<0.001) and associated with prevalent atrial fibrillation (OR=1.48 95%CI 1.07-2.05, p=0.019, adjusted for age, sex, systolic blood pressure and NT-proBNP)

**Figure 4.**
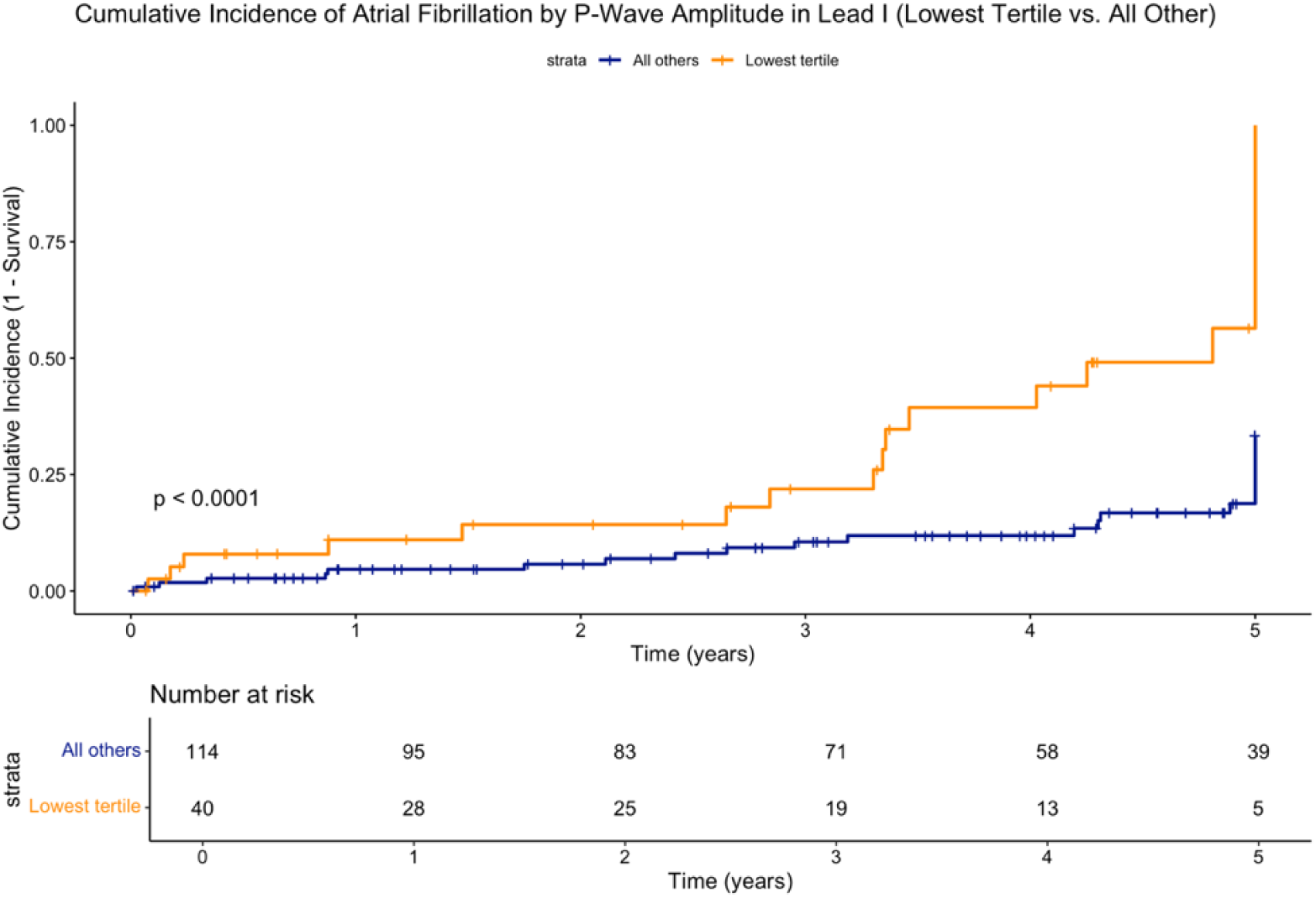
Cumulative incidence of AF by P-wave amplitude in lead I (≤ 0.04 mV) vs all other)..

**Table 3.** Cox regression analysis for associations between fibrotic biomarkers and incident AF (n=102, including 33 patients with incident AF during follow up).

| Biomarker | Model 1 |  |  | Model 2 |  |  | Model 3 |  |  |
| --- | --- | --- | --- | --- | --- | --- | --- | --- | --- |
|  | HR | 95%CI | p-value | HR | 95%CI | p-value | HR | 95%CI | p-value |
| TIMP-4 | **1.83** | **1.15–2.90** | **0.011** | **2.05** | **1.21–3.47** | **0.008** | **2.06** | **1.21–3.48** | **0.007** |
| MMP-2 | **2.18** | **1.21–3.93** | **0.009** | **2.09** | **1.03–4.21** | **0.041** | **2.09** | **1.01–4.30** | **0.046** |
| MMP-3 | **1.44** | **1.11–1.86** | **0.005** | **1.48** | **1.12–1.95** | **0.006** | **1.48** | **1.12–1.96** | **0.007** |
| MMP-9 | 0.92 | 0.64–1.32 | 0.640 | 1.23 | 0.72–1.58 | 0.755 | 1.06 | 0.72–1.56 | 0.768 |
| ST-2 | **1.37** | **1.00–1.86** | **0.048** | **1.64** | **1.17–2.28** | **0.004** | **1.66** | **1.19–2.34** | **0.003** |
| GDF-15 | **2.60** | **1.69–4.30** | **<0.001** | **2.20** | **1.36–3.56** | **0.001** | **2.26** | **1.38–3.70** | **0.001** |
| Gal-3 | 1.85 | 0.93–3.69 | 0.080 | 2.07 | 0.98–4.37 | 0.056 | **2.14** | **1.01–4.53** | **0.048** |

TIMP-4, Metalloproteinase inhibitor 4. Matrix metalloproteinase 2, 3, and 9 (MMP-2, MMP-3, and MMP-9, respectively). ST-2, suppression of tumorigenicity 2. GDF-15. Growth/differentiation factor 15. GAL-3, galectin-3.
HR: Hazard ratio
CI: Confidence interval.
Model 1: Unadjusted
Model 1: Adjusted for Age, Sex
Model 2: Adjusted for Age, Sex and NT-proBNP

**Table 4.** Cox regression analysis for associations between p-wave indices and incident AF (n=153, including 41 patients with incident AF during follow up).

| P-wave indices | Model 1 |  |  | Model 2 |  |  | Model 3 |  |  |
| --- | --- | --- | --- | --- | --- | --- | --- | --- | --- |
|  | HR | 95%CI | p-value | HR | 95%CI | p-value | HR | 95%CI | p-value |
| P-wave duration $\geq 120$ ms | 0.79 | 0.42–1.51 | 0.476 | 0.89 | 0.46–1.72 | 0.773 | 0.86 | 0.44–1.66 | 0.649 |
| PTF- V1 $\geq 0.04$ mm·s | 0.95 | 0.50–1.81 | 0.875 | 1.22 | 0.63–2.36 | 0.560 | 1.24 | 0.64–2.41 | 0.521 |
| P-wave axis $< 0^\circ$ | **8.17** | **2.26–29.6** | **0.009** | **5.17** | **1.39–19.2** | **0.014** | **4.74** | **1.26–17.7** | **0.021** |
| P-wave axis $> 75^\circ$ | 1.13 | 0.44–2.94 | 0.797 | 1.40 | 0.54–3.65 | 0.486 | 0.90 | 0.55–3.74 | 0.461 |
| P-amplitude in lead I (lowest tertile, $\leq 0.040$ mV) | **2.44** | **2.23–4.56** | **<0.005** | 1.93 | 0.99–3.76 | 0.054 | **2.09** | **1.05–4.14** | **0.036** |
| Biphasic P-wave (lead aVF) | 1.84 | 0.65–5.18 | 0.252 | 1.65 | 0.57–4.78 | 0.360 | 1.75 | 0.60–5.07 | 0.304 |
| Biphasic P-wave (lead II) | 1.22 | 0.63–2.37 | 0.556 | 1.23 | 0.63–2.39 | 0.540 | 1.36 | 0.69–2.67 | 0.378 |
| Advanced IAB | 0.95 | 0.13–6.91 | 0.959 | 0.85 | 0.12–6.27 | 0.874 | 1.15 | 0.15–8.75 | 0.895 |
| Partial IAB | 0.77 | 0.39–1.51 | 0.446 | 0.87 | 0.42–1.71 | 0.691 | 0.80 | 0.40–1.60 | 0.524 |

IAB – interatrial block; PTF-V1 – P-wave terminal force in lead V1
HR: Hazard ratio
CI: Confidence interval
Model 1: Unadjusted
Model 1: Adjusted for Age, Sex
Model 2: Adjusted for Age, Sex, SBP, and NT-proBNP

## Discussion

To the best of our knowledge, this is the first prospective long-term follow-up study that demonstrates significant associations between the biomarkers linked to myocardial fibrosis, electrocardiographic indices of atrial abnormality and incident AF in patients admitted for acute HF. The strongest association was observed for GDF-15, MMP-2 and TIMP-4, which demonstrated significant association with both the prevalent AF at enrolment and incident AF during follow-up. Moreover, GDF-15 and MMP-2 significantly correlated to the reduced P-wave amplitude, which is a novel finding that further supports the use of P-wave amplitude as an indicator of atrial substrate associated with AF. In addition to the reduced P-wave amplitude, abnormal P-wave axis <0° was significantly associated with incident AF. These findings underscore the importance of assessing P-wave characteristics in standard electrocardiograms for early identification of individuals at elevated risk of AF. Incorporating PWI together with biomarkers enhance AF risk stratification and inform preventive strategies.

### Interplay between atrial fibrillation heart failure and electrocardiogram properties with potential mechanism

The complex interplay between AF and HF involves structural, electrical, and functional changes that perpetuate each other’s progression while contributing to adverse clinical outcomes and creating a self-reinforcing vicious cycle [20]. Hemodynamic overload due to HF increases pressure and volume in the atria, leading to myocyte death and atrial remodeling. Additionally, mechanical stretch of the atria modulates mechano-electric feedback, impairing electrical signaling and promoting AF [20, 21]. Ultimately, myocyte death, combined with prolonged mechanical stretch, initiates atrial fibrosis, creating conduction heterogeneity that disrupts the normal electrical pathways and facilitates triggered activity and reentry - a hallmark of AF [22, 23]. Electrocardiograms allow for the estimation of atrial conduction properties, and PWI reflect delays and heterogeneity in atrial conduction caused by fibrosis. These indices serve as non-invasive markers for diagnosing atrial cardiomyopathy and assessing AF risk [24]. For instance, in the Framingham and ARIC studies, a P-wave duration >120 ms was significantly associated with AF over a 10-year follow-up. However, the prevalence of HF in those populations was less than 1% [4]. In a meta-analysis, the ARIC study reported that an abnormal P-wave axis (<0° or >75°) significantly elevated AF risk [25]. Consistent with our findings where a P-wave axis less than 0° was significantly associated with incident AF, however the association with >75° was not observed in either the cross-sectional analysis of prevalent AF or the Cox regression of incident AF. Findings from the Malmö Preventive project indicated that neither P-wave duration, PR interval, nor PTF-V1 was associated with incident AF in an elderly population, suggesting their limited value as risk indicators [26]. While PWI have previously been linked to atrial remodeling and incident AF in healthy individuals [27], evidence in acute HF populations is scare. A pooled analysis of over 25, 000 individuals from four prospective population cohorts demonstrated that multiple P-wave abnormalities on ECG are associated with increased lifetime risk of AF [28]. Participants with ≥4 PWI abnormalities developed AF roughly 4.6 years earlier than those without abnormalities [28]. In our cohort, a substantial proportion of the individuals exhibited at least 2 P-wave abnormalities, aligning the with recent findings and this may explain the high arrhythmia burden observed in advance heart failure. In advanced heart failure with reduced ejection fraction (HFrEF) requiring cardiac resynchronization therapy (CRT), IAB has shown significant association with new-onset AF [29]. In our cohort, advanced IAB was significantly associated with the prevalent AF, however the number of patients with this ECG phenomenon was too few among those who did not have AF history at baseline (n=5), which is likely the reason why this association could not be demonstrated in the analysis of incident AF. Our findings further support the role of the low P-wave amplitude in lead I as an indicator of atrial cardiomyopathy associated with AF development. It has been shown that low P-wave amplitude (<0.1 mV) in lead I was related to displaced inter-atrial conduction pattern and independently predicted recurrence of paroxysmal AF in patients undergoing radiofrequency catheter ablation [30]. We advanced these observations by demonstrating that low P-wave amplitude in lead I was significantly associated not only with prevalent AF but also with new onset AF in patients who did not have AF history at admission with acute HF. Significant correlation with several biomarkers that have established link to development of myocardial fibrosis (such as GDP-15 and MMP-2), which we observed in our cohort, further support the pathophysiological link between low P-wave amplitude and AF.

### Biomarkers relation to atrial fibrillation in heart failure

We have previously demonstrated in the that increased plasma levels of TIMP-4, GDF-15, and ST-2 are associated with the prevalence of AF[10]. In the current study, we expand upon these findings by showing that these proteins, along with Galectin-3, MMP-2, and MMP-3, are also associated with the incidence of AF. The association is likely explained by their involvement in fibrotic and remodeling processes within the myocardium and atrial tissue, contributing to the structural and electrical changes that predispose the participants to AF. **GDF-15,** a member of the TGF-β family and is upregulated in response to cellular stressors, including hypoxia, inflammation, and oxidative stress[31]. It has been identified as a significant biomarker for predicting mortality in conditions such as HF [32] and myocardial infarction[33], as well as for evaluating thrombosis risk in patients with AF [34]. To our knowledge, this study is the first to demonstrate a longitudinal association between GDF-15 levels and the onset of AF in an HF population. Given that GDF-15 levels correlate with the severity of HF[35], their association with incident AF observed in this study may reflect GDF-15’s ability to signal advanced stages of HF, which in turn heightens the risk of developing AF.

**Matrix metalloproteinases (MMPs**), specifically MMP-2 and MMP-9, are zinc-dependent endopeptidases essential for extracellular matrix remodeling, including in atrial tissue [36]. Our study identified increased MMP-2 and MMP-3 levels as significantly associated with incident AF, potentially due to the MMPs role in promoting atrial remodeling via fibrotic and inflammatory mechanisms, as well as its association with systemic stress responses that facilitate arrhythmogenesis [37].

**ST-2** is a member of the interleukin-1 receptor family[38] and together with interleukin-33 form a cardioprotective signal pathway and regulates the myocardial response to increased hemodynamic load and atrial stretch[15, 38, 39]. The finding that f ST2 is associated with incident AF raises the question of whatever elevated ST-2 levels are primarily driven by to high heart rate and elevated atrial pressure, or if it reflects underlying advanced HF.

**Galectin-3** is a galactoside-binding lectin involved in inflammatory and fibrosis responses and leads to the proliferation of fibroblasts, collagen disposition following cardiac fibrosis, and myocyte disruption[40]. It has been shown to contribute to adverse structural and electrophysiological remodeling in the atria [41], which may explain our findings demonstrating that elevated levels are associated with incident AF.

### Strength and Limitations

Our study is an observational study with an elderly population in different HF stages, with several other cardiovascular co-morbidities, which limits generalizability. However, the use of limited exclusion criteria ensures that the study population closely mirrors a real-world representation of individuals with HF and thereby strengthening its generalizability and real-data relevance. This study includes comprehensive definition and diagnosis of AF but relatively small sample size. Additionally, there is a possibility that not all cases of AF have been recorded due to the lack of prolonged rhythm monitoring but also due to the sometimes paroxysmal nature of this arrythmia. Further, not all patients had available biomarker data. Finally, during the follow-up, there is a possibility of non-identified cases new-onset of AF.

## Conclusion

This study showed that low P-wave amplitude in lead I and abnormal P-wave axis (<0 degree) was strongly associated with incident AF. In this prospective long-term follow-up study of patients hospitalized for acute heart failure, six out of seven biomarkers linked to cardiac fibrosis showed significant associations with the occurrence of AF. While the exact mechanisms of biomarkers remain unclear, their direct involvement in myocardial remodeling processes and capacity to detect molecular and cellular changes in advanced HF provide a plausible explanation for their predictive advantage. Biomarkers such as GDF-15 and MMP-2 has strong predictive value for the development of AF based upon its correlation with low P-wave amplitude in patients with advance heart failure cohort.

## Non-standard Abbreviations and Acronyms

AF: Atrial Fibrillation
PWI: P-wave Indices
PWD: P-wave Duration
PTFV1: P-wave Terminal Force in lead V1
P-axis: P-wave Axis
IAB: Interatrial Block
HF: Heart Failure
HARVEST: HeARt and Brain Falure inVESTigation
ECG: Electrocardiogram
MUSE: (Proprietary ECG database/software by GE Healthcare)
TIMP-4: Tissue Inhibitor of Metalloproteinase 4
ST-2: Suppression of Tumorigenicity 2
Gal-3: Galectin-3
GDF-15: Growth/Differentiation Factor 15
MMP-2/3/9: Matrix Metalloproteinase 2, 3, and 9
CVD: Cardiovascular Disease
PEA: Proximity Extension Assay
SBP: Systolic Blood Pressure
DBP: Diastolic Blood Pressure
BMI: Body Mass Index
NT-proBNP: N-terminal pro-B-type Natriuretic Peptide
EQUALIS: External Quality Assurance in Laboratory Medicine in Sweden
ANOVA: Analysis of Variance
FDR: False Discovery Rate
HR: Hazard Ratio
CI: Confidence Interval
ARIC: Atherosclerosis Risk in Communities (Study)
CRT: Cardiac Resynchronization Therapy
HFrEF: Heart Failure with Reduced Ejection Fraction

## Data Availability

The data that support the findings of this study are available from the corresponding author upon reasonable request.

## Acknowledgements

We thank the research nurses Hjördis Jernhed and Dina Chatziapostolou for valuable contributions. The Knut and Alice Wallenberg foundation is acknowledged for generous support.

